# DEVELOPING AND VALIDATING COVID-19 ADVERSE OUTCOME RISK PREDICTION MODELS FROM A BI-NATIONAL EUROPEAN COHORT OF 5594 PATIENTS

**DOI:** 10.1101/2020.10.06.20207209

**Authors:** Espen Jimenez-Solem, Tonny S Petersen, Casper Hansen, Christian Hansen, Christina Lioma, Christian Igel, Wouter Boomsma, Oswin Krause, Stephan Lorenzen, Raghavendra Selvan, Janne Petersen, Martin Erik Nyeland, Mikkel Zöllner Ankarfeldt, Gert Mehl Virenfeldt, Matilde Winther-Jensen, Allan Linneberg, Mostafa Mediphour Ghazi, Nicki Detlefsen, Andreas Lauritzen, Abraham George Smith, Marleen de Bruijne, Bulat Ibragimov, Jens Petersen, Martin Lillholm, Jon Middleton, Stine Hasling Mogensen, Hans-Christian Thorsen-Meyer, Anders Perner, Marie Helleberg, Benjamin Skov Kaas-Hansen, Mikkel Bonde, Alexander Bonde, Akshay Pai, Mads Nielsen, Martin Sillesen

## Abstract

**Background:** Patients with severe COVID-19 have overwhelmed healthcare systems worldwide. We hypothesized that Machine Learning (ML) models could be used to predict risks at different stages of management (at diagnosis, hospital admission and ICU admission) and thereby provide insights into drivers and prognostic markers of disease progression and death.

**Methods:** From a cohort of approx. 2.6 million citizens in the two regions of Denmark, SARS-CoV-2 PCR tests were performed on subjects suspected for COVID-19 disease; 3944 cases had at least one positive test and were subjected to further analysis. A cohort of SARS-CoV-2 positive cases from the United Kingdom Biobank was used for external validation.

**Findings:** The ML models predicted the risk of death (Receiver Operation Characteristics – Area Under the Curve, ROC-AUC) of 0.904 at diagnosis, 0.818, at hospital admission and 0.723 at Intensive Care Unit (ICU) admission. Similar metrics were achieved for predicted risks of hospital and ICU admission and use of mechanical ventilation. We identified some common risk factors, including age, body mass index (BMI) and hypertension as driving factors, although the top risk features shifted towards markers of shock and organ dysfunction in ICU patients. The external validation indicated fair predictive performance for mortality prediction, but suboptimal performance for predicting ICU admission.

**Interpretation:** ML may be used to identify drivers of progression to more severe disease and for prognostication patients in patients with COVID-19. Prognostic features included age, BMI and hypertension, although markers of shock and organ dysfunction became more important in more severe cases.

We provide access to an online risk calculator based on these findings.

**Funding:** The study was funded by grants from the Novo Nordisk Foundation to MS (#NNF20SA0062879 and #NNF19OC0055183) and MN (#NNF20SA0062879). The foundation took no part in project design, data handling and manuscript preparation.

## INTRODUCTION

The COVID-19 pandemic has put severe strains on hospital systems around the world. As of October 1st, 2020, the World Health Organization (WHO) estimated that more than 34 million patients are affected worldwide, and that the pandemic is the direct cause of more than 1 million deaths - a number that will likely rise as the pandemic progress.

The unknown clinical features coupled with the speed of viral spreading creates an unfortunate situation where health care providers are lacking important diagnostic adjuncts such as accurate prediction models and data-driven insights into the drivers of disease progression.

Several studies have now proposed prediction models based on a variety of clinical features. The majority of these are, however, trained and validated on national datasets from hospital admitted COVID-19 patients. While these may be of value locally, whether the classification ability transfers to other healthcare systems is questionable. As such, a recent review of Machine Learning (ML) models^1^, aimed at risk prediction in SARS-CoV-2 positive patients, found that the majority of studies were constructed on Chinese data with a high risk of bias as assessed by the Prediction model Risk of Bias Assessment tool (PROBAST)^2^. Furthermore, models often utilize data from hospital admitted SARS-CoV-2 positive patients only, which may skew results due to the lack of data from patients with milder disease trajectories.

Finally, models are often developed for predictions at hospital admission, providing little insight into the effects of in-hospital management. This, in turn, hinders identification of optimal biomarkers and prognostic features of adverse outcomes, as these may change as the patient advance through the trajectory of the disease.

The objective of this study is to construct and validate an ML model for SARS-CoV-2 adverse outcome risk prediction on a European dataset from Denmark, with external validation in a United Kingdom (UK) dataset. Secondly, we seek to use the constructed models for identification of disease risk factors, including comorbidities, biomarkers and vital signs as the patient moves through the disease trajectory. As such, we seek to identify important clinical predictors of adverse outcomes at four timepoints or timeframes, based on accumulating available data: On diagnoses, on hospital admission (where applicable) and immediately before and after admission to the intensive care unit (ICU9), where applicable. Finally, we sought to create an online prediction tool to support rapid risk assessment upon COVID-19 diagnosis based on the most relevant data points.

We hypothesized that ML can be leveraged to provide accurate outcome predictions for COVID-19 patients, and that including accumulating datapoints from available sources from Electronic Health Record (EHR) repositories in a combined model improves risk prediction as well as identification of relevant disease drivers at specific time points.

## METHODS

The study was approved by the relevant legal and ethics boards. These included the Danish Patient Safety Authority (Styrelsen for Patientsikkerhed, approval #31-1521-257) and the Danish Data Protection Agency (Datatilsynet, approval #P-2020-320) as well as the UK Biobank (Application ID #60861) COVID-19 cohort. Under Danish law, approval from these agencies are required for access to and handling of patient sensitive data, including EHR records. Legal approval for the study was furthermore obtained from the Danish Capital Region (Region Hovedstaden).

We conducted a prospective study by including all individuals undergoing a SARS-CoV-2 test (nasal and/or pharyngeal swap subjected to Real-Time Polymerase Chain Reaction testing) in the Capital and Zealand Regions (approximately 2.6 million citizens) of Denmark between March 1^st^, 2020 and June 16^th^ 2020. Data inclusion was censored on June 16^th^. Patients were identified through their Central Person Registry (CPR) number, a unique numerical combination given to every Danish citizen, enabling linking of electronic health records (EHRs) with nationwide medical registry data.

During the study period, all SARS-CoV-2 tests were performed at regional hospitals. Patients were referred for testing based on presence of symptoms albeit test strategies shifted towards the end of the inclusion period to include a wider screening indication.

For cases with at least one positive tests, we extracted data from the bi-regional EHR system, including demographics, comorbidities and prescription medication. In-hospital data included laboratory results and vital signs.

Supplementary Table S1 lists extracted comorbidities with their definitions, Supplementary Table S2 extracted laboratory values, and Supplementary Table S3 extracted temporal features (vital signs).

For the purpose of external validation of the ML models, we extracted data from the UK biobank COVID-19 cohort. The UK biobank contains detailed healthcare information on 500.000 UK citizens, of which 1650 have been tested SARS-CoV-2 positive. This cohort has recently been made available for the purpose of COVID-19 research by the UK biobank consortium^3^.

### Prediction models

ML models were trained and validated on the Danish dataset. A subset of models sharing identical data fields (e.g. age, comorbidities etc.) between the Danish and UK cohorts were subsequently externally validated on the UK biobank dataset.

We constructed ML prediction models by including available data for patients up to and including the selected time frames or time points. These time frames or points were

- At time of SARS-CoV-2 positivity (all patients, Diagnosis model)
- The first 12 hours of hospital admission (Admission model)
- 12 hours up to ICU admission (Pre-ICU model)
- 12 hours after ICU admission (Post-ICU model).

Models were trained to predict one of four events, where applicable:

- Hospital admission (SARS-CoV-2 positive patients)
- ICU admission (Diagnosed and hospital admitted patients)
- Mechanical ventilation (Diagnosed, hospital and ICU admitted patients)
- Death (all patients)

For each task, we trained with different feature sets to study how incrementally adding data affects model performance as well as to gain insight into drivers of disease progression:

- Base models: Age, sex and body mass index (BMI).
- Comorbidities: Base model and comorbidities (Table 1 and Supplementary Table S1)
- Temporal features: Comorbidities model and temporal features (Supplementary Table S3)
- In-hospital laboratory tests: Temporal features model and in-hospital lab tests (Supplementary Table S2).

**Table 1:** Demographic information on the group of SARS-CoV-2 positive patients, including information on pre-existing comorbidities. Supplementary Table S2 holds information on diagnoses codes included in the individual comorbidity classifications. The table presents information on the full cohort (admitted and non-admitted SARS-CoV-2 positive patients) as well as subgroups admitted to a hospital and Intensive Care Unit (ICU) respectively. Furthermore, differential demographics between survivors and non-survivors (in-hospital mortality) is presented. Continuous variables are presented as medians with (interquartile range) **p<0.001 when subgroups are compared (e.g. hospitalized vs. non-hospitalized, ICU vs. non-ICU, survivors vs. non- survivors). COPD: Chronic Obstructive Pulmonary Disease.

|  | All SARS-CoV-2 patients (n=3944) | Non-hospitalized (n=2585) | Hospitalized patients (n=1359) | Hospitalized patients without ICU admission (n=1178) | Hospitalized patients with ICU admission (n=181) | Survivors (n=3620) | Non-survivors (n=324) |
| --- | --- | --- | --- | --- | --- | --- | --- |
| Body Mass Index | 25.9 (22.7-29.8) | 25.8 (22.6-29.7) | 26.0 (22.7-29.9) | 25.7 (22.6-29.4) | 27.3 (23.5-31.4) | 26.0 (22.7-29.9) | 25.2 (22.0-28.8) |
| Age | 53.0 (36.0-69.0) | 44.0 (31.0-58.0) | 70.0 (55.0-80.0)** | 71.0 (54.0-81.0) | 69.0 (58.0-75.0) | 50.0 (34.0-64.0) | 81.0 (73.0-87.0)** |
| Male Sex | 41.9 | 35.6 | 53.9** | 51.4 | 70.2** | 40.6 | 56.2** |
| Diabetes | 13.4 | 8.0 | 23.5** | 23.4 | 24.3 | 12.0 | 28.7** |
| Ischemic heart disease | 9.1 | 5.2 | 16.5** | 16.0 | 19.3 | 8.0 | 21.0** |
| Heart failure | 4.2 | 1.5 | 9.4** | 9.5 | 8.8 | 3.1 | 16.7** |
| Arrhythmia | 10.4 | 4.9 | 20.9** | 21.0 | 20.4 | 8.5 | 32.1** |
| Stroke | 6.7 | 3.1 | 13.5** | 14.1 | 9.9 | 5.7 | 17.9** |
| COPD or Asthma | 11.4 | 9.2 | 15.7** | 15.4 | 17.1 | 10.9 | 17.0** |
| Sleep apnoea | 0.0 | 0.0 | 0.0 | 0.0 | 0.0 | 0.0 | 0.0 |
| Arthritis | 1.6 | 1.4 | 2.2 | 2.1 | 2.8 | 1.5 | 3.1 |
| Osteoporosis | 6.0 | 2.9 | 12.1** | 12.0 | 12.7 | 4.9 | 18.5** |
| Dementia | 2.9 | 1.3 | 5.9** | 6.7 | 0.6** | 1.8 | 14.8** |
| Severe mental disorder | 1.7 | 1.5 | 2.0 | 2.1 | 1.1 | 1.5 | 4.3** |
| Immunodeficiencies | 0.4 | 0.3 | 0.5 | 0.5 | 0.6 | 0.4 | 0.6 |
| Neurological manifestations | 18.1 | 15.0 | 24.1** | 25.1 | 17.1 | 16.8 | 33.0** |
| Cancer | 10.3 | 6.3 | 18.0** | 17.8 | 18.8 | 8.8 | 27.5** |
| Chronic kidney failure | 2.4 | 0.7 | 5.7** | 5.6 | 6.6 | 1.8 | 9.6** |
| Dialysis | 0.5 | 0.1 | 1.3** | 1.3 | 1.1 | 0.4 | 1.5 |
| Hypertension | 30.1 | 18.3 | 52.5** | 48.6 | 78.5** | 27.0 | 65.1** |

For the purpose of external validation, data points were available in the UK biobank matching those of the base and comorbidities models. In-hospital models could not be externally validated due to lack of availability of these data points in the UK biobank.

### ML models

We used *random forests (RFs)*^*4*^, implemented in the open-source machine learning library *scikit-learn*^*5*^. Because each individual tree is trained on a bootstrap sample, there is training data that can be used to compute the *out-of-bag (OOB) error*, an estimate of the RF’s performance.

All models were evaluated on the Danish set using 5-fold cross-validation. The folds were stratified to ensure that the splits are representative of the full cohort. For each split, we conducted grid search on the available training data fold to tune the hyperparameters of the RF models for each prediction task. The OOB error served as the selection criterion. We varied the number of decision trees in the ensemble in 100, 500, 1000 and the maximum depth of the individual decision trees in 3,4,7,9,11. When splitting a node, all input features were checked.

For computing the Receiver Operating Characteristics Area Under the Curve (ROC-AUC) and precision/recall AUC (PR-AUC), the outputs for all test folds were combined, resulting in predictions for the entire data set.

For evaluation on the UK data, models were trained on the entire Danish data set. As before, for each model, a grid search based on the OOB error was performed on the same parameter grid. Each model was then evaluated on the entire UK cohort.

Post-hoc analysis of the use of the predictive variables across all decision trees in the RF allows us to derive a measure of feature importance. Feature importance was calculated by the *mean decrease in impurity* (MDI). The measure takes into account how often a feature is used when classifying the training data points and how well it splits the training data points when being used. For each predictive variable the importance was computed as the mean over the feature importance for each fold. The top-10 or top-20 (depending on the model) were extracted and visualized. The correlations of these features were then computed across the entire dataset.

### Missing data

Missing data was considered missing at random.

Percentages of available data in the Danish cohort are presented in supplementary Table S4. Missing values for BMI were imputed by using k-nearest neighbour imputation using age and sex^6^, with k=100. Other missing data points were set to “not available” for the purpose of ML modelling and deleted by case wise deletion for group comparisons.

### Data presentation and statistical testing

Continuous data is presented as medians (interquartile range) and compared using the Mann-Whitney U test. Categorical data is presented as percentages and compared using the Chi-square test.

ML model performances are presented as ROC-AUC for Positive and Negative predictive value and precision/recall. Model comparisons were performed by the deLong test^7^.

The p-values for comparisons of outcome groups are provided for reference only. As these comparisons are not part of the study hypotheses, p-values are presented without post-hoc correction for multiple testing and should be interpreted as such.

In addition, calibration curves are presented for the combined test folds and the external validation data. For each calibration plot, the predictions were grouped using quantile-based binning.

### Online models

The risk prediction model for SARS-CoV-2 positive patients admitted to hospital is available in an online version on objhttps://cope.science.ku.dk.

## RESULTS

A total of 3944 individuals had at least one positive SARS-CoV-2 test in the two Danish regions and were included in the study. These were supplemented by the 1650 patients from the UK biobank used for external model validation.

Among the Danish cases, 1359 (34.5%) required hospitalization, and 181 (4.6%) intensive care. A total of 324 patients (8.2%) died.

Demographics and comorbidities are summarized in Table 1, selected laboratory values and vital signs in Table 2. Demographics information for the UK biobank external validation cohort is presented in supplementary Table S5.

**Table 2:**
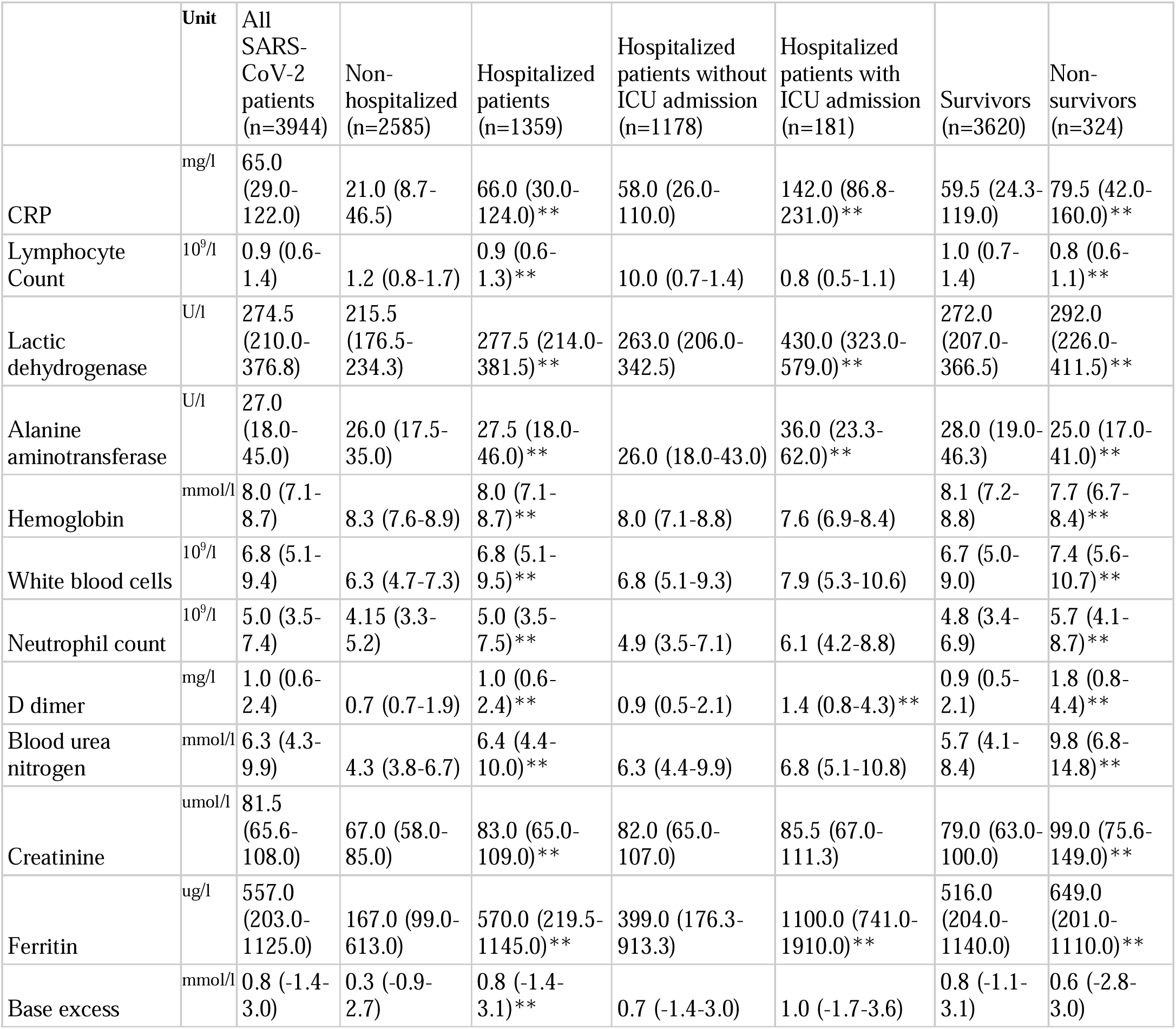
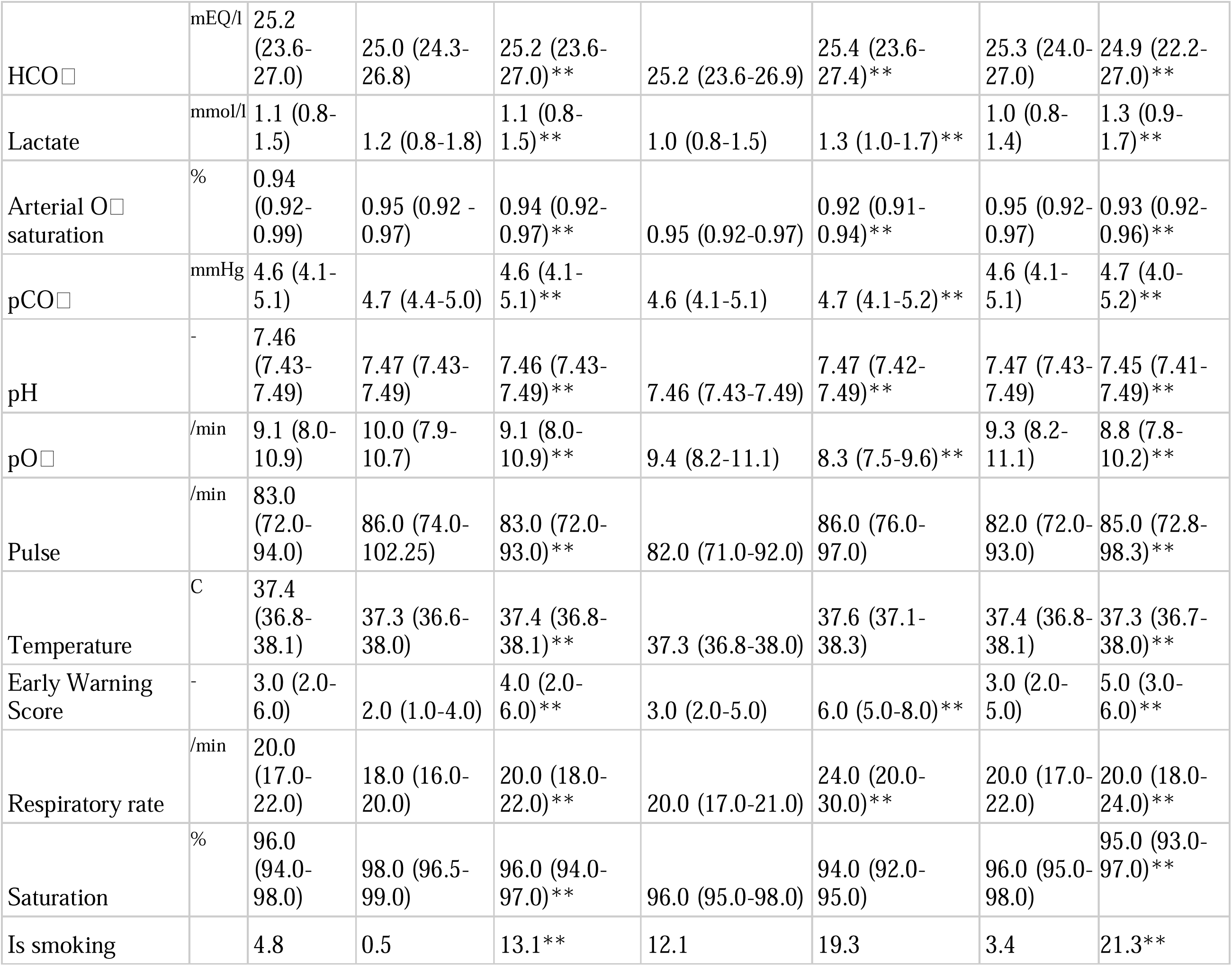
Temporal features (laboratory tests and vital signs). The table presents information on the full cohort (admitted and non-admitted SARS-CoV-2 positive patients) as well as subgroups admitted to a hospital and Intensive Care Unit (ICU) respectively. Furthermore, differential temporal features between survivors and non-survivors (in-hospital mortality) is presented. Continuous variables are presented as medians with (interquartile range) **p<0.001 when subgroups are compared (e.g. hospitalized vs. non-hospitalized, ICU vs. non-ICU, survivors vs. non- survivors).

|  | Unit | All SARS-CoV-2 patients (n=3944) | Non-hospitalized (n=2585) | Hospitalized patients (n=1359) | Hospitalized patients without ICU admission (n=1178) | Hospitalized patients with ICU admission (n=181) | Survivors (n=3620) | Non-survivors (n=324) |
| --- | --- | --- | --- | --- | --- | --- | --- | --- |
| CRP | mg/l | 65.0 (29.0-122.0) | 21.0 (8.7-46.5) | 66.0 (30.0-124.0)** | 58.0 (26.0-110.0) | 142.0 (86.8-231.0)** | 59.5 (24.3-119.0) | 79.5 (42.0-160.0)** |
| Lymphocyte Count | 10<sup>9</sup>/l | 0.9 (0.6-1.4) | 1.2 (0.8-1.7) | 0.9 (0.6-1.3)** | 10.0 (0.7-1.4) | 0.8 (0.5-1.1) | 1.0 (0.7-1.4) | 0.8 (0.6-1.1)** |
| Lactic dehydrogenase | U/l | 274.5 (210.0-376.8) | 215.5 (176.5-234.3) | 277.5 (214.0-381.5)** | 263.0 (206.0-342.5) | 430.0 (323.0-579.0)** | 272.0 (207.0-366.5) | 292.0 (226.0-411.5)** |
| Alanine aminotransferase | U/l | 27.0 (18.0-45.0) | 26.0 (17.5-35.0) | 27.5 (18.0-46.0)** | 26.0 (18.0-43.0) | 36.0 (23.3-62.0)** | 28.0 (19.0-46.3) | 25.0 (17.0-41.0)** |
| Hemoglobin | mmol/l | 8.0 (7.1-8.7) | 8.3 (7.6-8.9) | 8.0 (7.1-8.7)** | 8.0 (7.1-8.8) | 7.6 (6.9-8.4) | 8.1 (7.2-8.8) | 7.7 (6.7-8.4)** |
| White blood cells | 10<sup>9</sup>/l | 6.8 (5.1-9.4) | 6.3 (4.7-7.3) | 6.8 (5.1-9.5)** | 6.8 (5.1-9.3) | 7.9 (5.3-10.6) | 6.7 (5.0-9.0) | 7.4 (5.6-10.7)** |
| Neutrophil count | 10<sup>9</sup>/l | 5.0 (3.5-7.4) | 4.15 (3.3-5.2) | 5.0 (3.5-7.5)** | 4.9 (3.5-7.1) | 6.1 (4.2-8.8) | 4.8 (3.4-6.9) | 5.7 (4.1-8.7)** |
| D dimer | mg/l | 1.0 (0.6-2.4) | 0.7 (0.7-1.9) | 1.0 (0.6-2.4)** | 0.9 (0.5-2.1) | 1.4 (0.8-4.3)** | 0.9 (0.5-2.1) | 1.8 (0.8-4.4)** |
| Blood urea nitrogen | mmol/l | 6.3 (4.3-9.9) | 4.3 (3.8-6.7) | 6.4 (4.4-10.0)** | 6.3 (4.4-9.9) | 6.8 (5.1-10.8) | 5.7 (4.1-8.4) | 9.8 (6.8-14.8)** |
| Creatinine | umol/l | 81.5 (65.6-108.0) | 67.0 (58.0-85.0) | 83.0 (65.0-109.0)** | 82.0 (65.0-107.0) | 85.5 (67.0-111.3) | 79.0 (63.0-100.0) | 99.0 (75.6-149.0)** |
| Ferritin | ug/l | 557.0 (203.0-1125.0) | 167.0 (99.0-613.0) | 570.0 (219.5-1145.0)** | 399.0 (176.3-913.3) | 1100.0 (741.0-1910.0)** | 516.0 (204.0-1140.0) | 649.0 (201.0-1110.0)** |
| Base excess | mmol/l | 0.8 (-1.4-3.0) | 0.3 (-0.9-2.7) | 0.8 (-1.4-3.1)** | 0.7 (-1.4-3.0) | 1.0 (-1.7-3.6) | 0.8 (-1.1-3.1) | 0.6 (-2.8-3.0) |
| HCO□ | mEQ/l | 25.2<br>(23.6-27.0) | 25.0 (24.3-26.8) | 25.2 (23.6-27.0)** | 25.2 (23.6-26.9) | 25.4 (23.6-27.4)** | 25.3 (24.0-27.0) | 24.9 (22.2-27.0)** |
| Lactate | mmol/l | 1.1 (0.8-1.5) | 1.2 (0.8-1.8) | 1.1 (0.8-1.5)** | 1.0 (0.8-1.5) | 1.3 (1.0-1.7)** | 1.0 (0.8-1.4) | 1.3 (0.9-1.7)** |
| Arterial O□ saturation | % | 0.94<br>(0.92-0.99) | 0.95 (0.92 - 0.97) | 0.94 (0.92-0.97)** | 0.95 (0.92-0.97) | 0.92 (0.91-0.94)** | 0.95 (0.92-0.97) | 0.93 (0.92-0.96)** |
| pCO□ | mmHg | 4.6 (4.1-5.1) | 4.7 (4.4-5.0) | 4.6 (4.1-5.1)** | 4.6 (4.1-5.1) | 4.7 (4.1-5.2)** | 4.6 (4.1-5.1) | 4.7 (4.0-5.2)** |
| pH | - | 7.46<br>(7.43-7.49) | 7.47 (7.43-7.49) | 7.46 (7.43-7.49)** | 7.46 (7.43-7.49) | 7.47 (7.42-7.49)** | 7.47 (7.43-7.49) | 7.45 (7.41-7.49)** |
| pO□ | /min | 9.1 (8.0-10.9) | 10.0 (7.9-10.7) | 9.1 (8.0-10.9)** | 9.4 (8.2-11.1) | 8.3 (7.5-9.6)** | 9.3 (8.2-11.1) | 8.8 (7.8-10.2)** |
| Pulse | /min | 83.0<br>(72.0-94.0) | 86.0 (74.0-102.25) | 83.0 (72.0-93.0)** | 82.0 (71.0-92.0) | 86.0 (76.0-97.0) | 82.0 (72.0-93.0) | 85.0 (72.8-98.3)** |
| Temperature | C | 37.4<br>(36.8-38.1) | 37.3 (36.6-38.0) | 37.4 (36.8-38.1)** | 37.3 (36.8-38.0) | 37.6 (37.1-38.3) | 37.4 (36.8-38.1) | 37.3 (36.7-38.0)** |
| Early Warning Score | - | 3.0 (2.0-6.0) | 2.0 (1.0-4.0) | 4.0 (2.0-6.0)** | 3.0 (2.0-5.0) | 6.0 (5.0-8.0)** | 3.0 (2.0-5.0) | 5.0 (3.0-6.0)** |
| Respiratory rate | /min | 20.0<br>(17.0-22.0) | 18.0 (16.0-20.0) | 20.0 (18.0-22.0)** | 20.0 (17.0-21.0) | 24.0 (20.0-30.0)** | 20.0 (17.0-22.0) | 20.0 (18.0-24.0)** |
| Saturation | % | 96.0<br>(94.0-98.0) | 98.0 (96.5-99.0) | 96.0 (94.0-97.0)** | 96.0 (95.0-98.0) | 94.0 (92.0-95.0) | 96.0 (95.0-98.0) | 95.0 (93.0-97.0)** |
| Is smoking |  | 4.8 | 0.5 | 13.1** | 12.1 | 19.3 | 3.4 | 21.3** |

When compared to non-hospitalized patients, hospital admitted patients were older and more likely to be male.

A number of comorbidities were overrepresented in the admitted subgroup. These included hypertension, diabetes, ischemic heart disease, heart failure, arrythmias, stroke, chronic obstructive pulmonary disease (COPD) or asthma, osteoporosis, neurological disease, cancer, chronic kidney failure and use of dialysis. Hospitalized patients were more likely to be smokers (Table 2).

For hospitalized patients requiring ICU admission vs. hospitalized patients without ICU admission, only male sex, Body Mass Index (BMI), dementia and hypertension differed between patients. ICU-admitted patients were furthermore more likely to be smokers. When compared to survivors, non-survivors were older and male (Table 1).

Non-survivors were furthermore more likely to suffer from hypertension, diabetes, ischemic heart disease, heart Failure, arrythmias, stroke, COPD or asthma, osteoporosis, Dementia, Mental disorders, Neurological disease, cancer, chronic kidney failure and use of dialysis.

When compared to non-admitted, admitted patients differed significantly in all measured values (Table 2). Among those hospitalised, those admitted to the ICU had derangements in many variables (Table 2). The same was observed for non-survivors compared with survivors (Table 2).

### ML models prediction

ML models are presented in Table 3 and graphically depicted in supplementary Figure S1 (Diagnosis model), Supplementary Figure S2 (Admission model), Supplementary Figure S3 (Pre-ICU model) and Supplementary Figure S4 (Post-ICU model).

**Table 3:** Main results from the prediction models. Predictions were performed with data available from four different time frames in the patient disease trajectories (left column): On diagnosis (Diagnoses model), On hospital admission and 12-hours into admission (Admission model), 12 hours leading up to Intensive Care Unit (ICU) admission (Pre-ICU model) and 12 hours after ICU admission (post-ICU model). Models were trained to predict risk of hospital admission, ICU admission, ventilator treatment and death (top row). All models were trained with incremental data, starting with age, gender and Body Mass Index, then adding comorbidity information, temporal features (e.g. vital signs) and finally by adding hospital laboratory tests where applicable. Please see supplementary tables S1 and S2 for data definitions. Performance metrics are presented as the Receiver Operating Characteristics Area Under the Curve (ROC-AUC) for True/False positive rates (TPR/FPR) and Precision/Recall (Pre/Rec). *Model is significantly (p<0.01) better than the base prediction model (Age+gender+Body Mass Index, BMI) #Model is significantly (p<0.01) better than the comorbidities model §Model is significantly (p<0.01) better than the temporal model --: Insufficient data available at the time point, or prediction irrelevant (e.g. predicting hospital admission for patients already in the ICU).

|  | Hospital admission |  | ICU admission |  | Ventilator treatment |  | Death |  |
| --- | --- | --- | --- | --- | --- | --- | --- | --- |
|  | TPR/FPR | Pre/Rec | TPR/FPR | Pre/Rec | TPR/FPR | Pre/Rec | TPR/FPR | Pre/Rec |
| **Diagnosis** |  |  |  |  |  |  |  |  |
| Age+Gender+BMI | 0.815 | 0.695 | 0.785 | 0.161 | 0.801 | 0.174 | 0.904 | 0.413 |
| +Comorbidities | 0.823 | 0.709 | 0.831* | 0.201 | 0.844* | 0.174 | 0.902 | 0.411 |
| +Temporal Features | -- | -- | -- | -- | -- | -- | -- | -- |
| +In-hospital Tests | -- | -- | -- | -- | -- | -- | -- | -- |
| **Admission** |  |  |  |  |  |  |  |  |
| Age+Gender+BMI | -- | -- | 0.678 | 0.221 | 0.699 | 0.206 | 0.787 | 0.434 |
| +Comorbidities | -- | -- | 0.727* | 0.271 | 0.718 | 0.224 | 0.798 | 0.443 |
| +Temporal Features | -- | -- | 0.72 | 0.261 | 0.722 | 0.254 | 0.797 | 0.449 |
| +In-hospital Tests | -- | -- | 0.792*#§ | 0.412 | 0.742 | 0.293 | 0.818* | 0.527 |
| **Pre-ICU** |  |  |  |  |  |  |  |  |
| Age+Gender+BMI | -- | -- | -- | -- | 0.587 | 0.878 | 0.731 | 0.58 |
| +Comorbidities | -- | -- | -- | -- | 0.552 | 0.842 | 0.702 | 0.536 |
| +Temporal Features | -- | -- | -- | -- | 0.593 | 0.885 | 0.724 | 0.549 |
| +In-hospital Tests | -- | -- | -- | -- | 0.446 | 0.849 | 0.723 | 0.579 |
| **Post-ICU** |  |  |  |  |  |  |  |  |
| Age+Gender+BMI | -- | -- | -- | -- | 0.587 | 0.878 | 0.731 | 0.58 |
| +Comorbidities | -- | -- | -- | -- | 0.56 | 0.845 | 0.723 | 0.558 |
| +Temporal Features | -- | -- | -- | -- | 0.559 | 0.868 | 0.733 | 0.56 |
| +In-hospital Tests | -- | -- | -- | -- | 0.683 | 0.927 | 0.736 | 0.591 |

Base models deployed on the time of diagnosis were able to predict hospital admission with a ROC-AUC of 0.815, ICU admission 0.785, ventilator treatment 0.801 and death 0.904 (Table 3 and Supplementary Figure S1).

Adding information on patient comorbidities increased the predictive ability for all outcomes. Models deployed at hospital admission achieved ROC-AUC scores ranging from 0.678 to 0.818 for the selected outcomes (Table 3 and Supplementary Figure S2). Adding information on comorbidities, temporal features and hospital laboratory tests increased model performance for ICU admission, use of mechanical ventilation and death.

Models deployed pre- or post-ICU admission achieved ROC-AUC’s from 0.587 to 0.736 (Table 3 and Supplementary Figure S3 and Supplementary Figure S4).

The calibration curves (supplementary figures S7 and S8) show that the models are well calibrated when looking at all diagnosed subjects and at patients admitted to the hospital. When restricted to patients admitted to ICU, the calibration gets worse as expected due to smaller sample size. Ventilator treatment could not be predicted accurately, and the calibration curves reflect this.

External validation results (Supplementary Table S6) on UK data indicated an overall reduction in model classification ability. For diagnosed patients, ROC-AUCs were 0.664 for predicting hospital admission, 0.548 for predicting ICU admission and 0.724 for predicting mortality. Inspection of the calibration curves (Supplementary Figure S7) shows that the models are only slightly worse calibrated for the UK data, meaning that the model outputs approximately the correct probability for individual patients, despite the degradation in ROC-AUC.

As patients progressed through the disease severity trajectories, mortality prediction remained in the area of 0.625 – 0.674 (Supplementary Table S6).

### Detection of important features and drivers of disease progression

Results of the drivers of disease progression feature detection analysis for each of the selected timepoints are depicted in Figure 1 (diagnoses model) and Figure 2 (admission model) as well as Supplementary Figure S4 (pre-ICU model) and Supplementary Figure S5 (post-ICU model).

**Figure 1:**
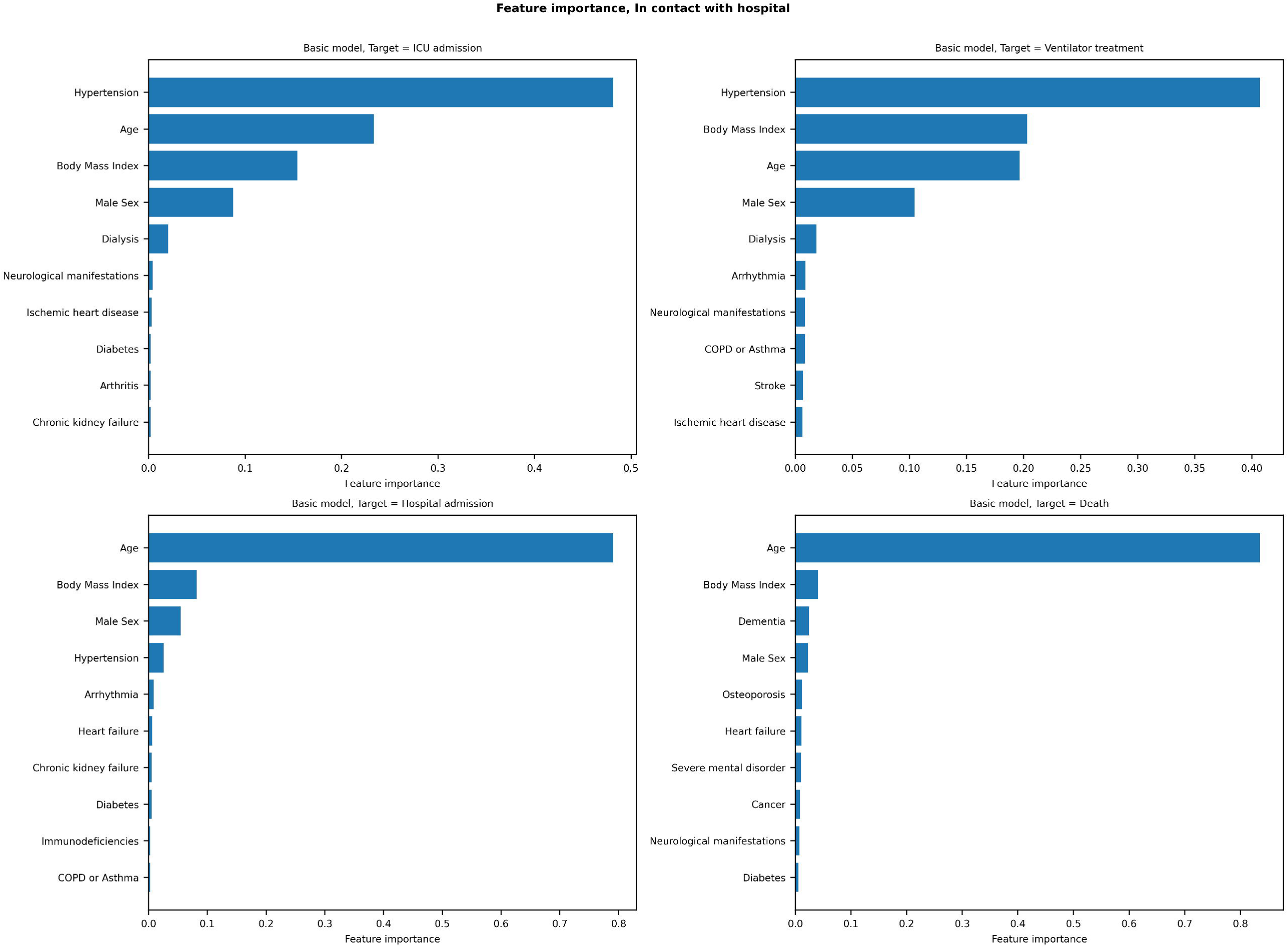
Feature importance for the basic (including age, sex, body mass index, and comorbidities) diagnosis models, predicting risk of intensive care admission (first row), hospital admission (second row), ventilator treatment (third row) and death (fourth row) on SARS-CoV-2 positivity.

**Figure 2:**
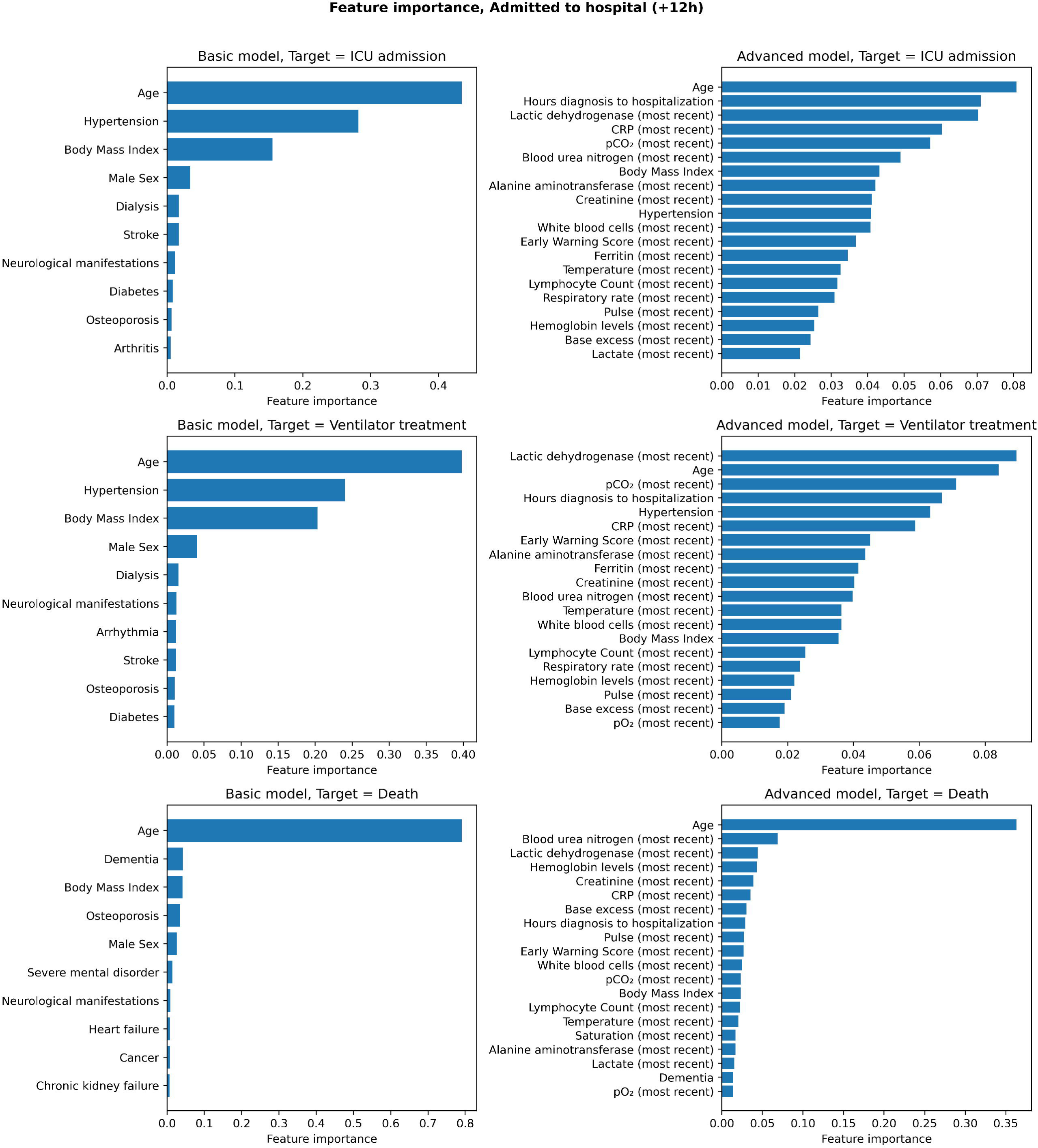
Feature importance for the basic (including age, sex, body mass index, and comorbidities) and advanced (all data) admission models, predicting risk of intensive care admission (first row), ventilator treatment (second row) and death (third row) on hospital admission.

For diagnosed patients (Figure 1), age and BMI were the most relevant features for risk of hospital admission and death, whereas hypertension was the most prominent risk feature for ICU admission and ventilator treatment.

Among comorbidities, hypertension was the most important feature for predicting ICU admission.

For admitted patients (Figure 2) the most relevant drivers of disease progression were age, BMI, hypertension and the presence of dementia. When the full dataset was analysed, lab tests indicating aspects of cell dysfunction (Lactic dehydrogenase, LDH), kidney dysfunction (Blood urea nitrogen and Creatinine), the inflammatory response (C-reactive protein, CRP), and liver damage (Alanine Aminotransferase, ALT) were identified as important prognostic markers for disease progression.

For ICU models (Supplementary Figure S5 and S6), relevant disease progression features again included age and BMI as well as comorbidities including hypertension, heart failure and neurological disease. When the full dataset was analysed, disease progression drivers included features reflecting insufficient respiration and metabolism (pulse oximetry oxygen saturation, pO2 values and slopes over time), as well as BUN and CRP levels.

## DISCUSSION

In this study, we analyse prognostic and factors associated with disease progression in 3944 SARS-CoV-2 positive patients by constructing an interpretable ML framework. In contrast to previous studies, these included diagnosed patients outside hospitals, and thus included the entire spectrum of SARS-CoV-2 positive patients in the 2.6 million regional population. Results indicate that by focusing on a limited number of demographic variables, including age, gender and BMI, it is possible to predict the risk of hospital and ICU admission, use of mechanical ventilation and death as early as at the time of diagnosis. Adding information on comorbidity to the model increased performance, indicating that these features play a prognostic role in the outcome of patients as they progress through the disease trajectory. As such, results from the ML feature detection indicate that comorbidity factors such as hypertension and diabetes are driving factors of adverse outcome, which is in line with reports from other cohort studies^8-10^. The role of hypertension is further underlined by reports indicating a role of the angiotensin converting enzyme 2 (ACE2) receptor as an entry point for the SARS-CoV2^11^. Whether COVID-19 interacts unfavourably with hypertension per se, or whether this risk is simply a manifestation of reduced tolerance to severe infection and hypoxia is currently debated^12^.

Furthermore, BMI was identified as a major feature of adverse outcome, as also reported by others^13-15^. Whether this is due to a reduced respiratory capacity or chronic impairment of the immune system through alterations in tumour necrosis factor and interferon secretion associated with obesity, is also currently debated^16^. Caution should, however, be taken when analysing these results, as the median observed differences between groups were minor and may not be clinically relevant. Furthermore, data imputation may have impacted on these results.

The addition of more data points, including temporal features and lab tests improved the model’s predictive value for hospitalized patients. Group comparisons indicated alterations of a plethora of laboratory tests for admitted patients, including features of immune activation and organ dysfunction. Interestingly, laboratory tests differed to a lesser extent between ICU and non-ICU patients, except for CRP levels, lymphocyte counts, LDH, ALT, neutrophil, D-dimer and ferritin levels as well as arterial blood gas values. As expected, ICU admitted patients had lower oxygen saturation and higher respiratory rates, likely reflecting the acute respiratory distress from COVID-19 pneumonia.

Feature analysis indicated that strong prognostic markers expectedly included CRP levels, but also markers of organ damage, including kidney injury (creatinine and blood urea nitrogen), liver injury (ALAT), cell damage (LDH), anaemia (haemoglobin levels) as well as ferritin levels. These, as well as vital signs and arterial blood gas values superseded many of the comorbidities in feature importance once the patient progressed through hospital and ICU admission. This again indicates that drivers and prognostic markers of adverse outcomes represent a dynamic field affected by the patient’s current point on the disease trajectory, and that differential values should be considered when risk-assessing COVID-19 patients depending on their current status (e.g. in hospital, in ICU etc.). A caveat is, however, that multiple comorbidities and advanced age may resulted in decisions by patients, relatives or clinicians limiting the use of life-support, and thus potentially precluding them from ICU admission and reducing the effect of comorbidities and age on model predictions. Kidney injury has previously been reported in patients with COVID-19^17,18^. Our finding that markers of kidney injury may be important at hospital admission supports the notion that COVID-19 associated kidney injury plays an important pathophysiological role.

The importance of LDH for COVID-19 patients has previously been reported in other ML^19^ as well as clinical studies^20^. These results are supported by the feature detection from this study, indicating that LDH levels serve as an important prognostic marker on hospital admission, although its value is superseded by other biomarkers when the patient advance to the ICU stage. As LDH can be seen as a general marker of cell and organ damage with a reported prognostic value for mortality in ICU patients^21^, these findings likely indicate a general organ affection associated with COVID-19 disease progression. Abnormal liver function tests, including ALAT, has previously been associated with COVID- 19 disease severity^22^. As such, reports have indicated the presence of elevated liver enzymes in both severe and non-severe COVID-19 cases^23^. Whether this is a function of viral infection, shock or a consequence of hepatotoxic treatments deployed during treatment is still not clear^24^.

Ferritin levels have previously been associated with COVID-19^25^, presumably due to its role in immunomodulation and association with the cytokine storm response seen in critical illness^26^.

Taken together, the feature importance of laboratory tests indicating affection of several organ systems indicates that COVID-19 disease severity follows a predictable pattern characterized by multi-organ affection (albeit not always dysfunction), which is in line with previous findings^27^.

Once patients progress to the ICU stage, feature detection indicated a switch towards vital signs and biomarkers indicating that the severity of respiratory failure, shock and inflammatory markers were the most important features of risk of death (Supplementary Figure S5 and Supplementary Figure S6).

When the feature importance of all models is analysed, the results indicate that COVID-19 outcomes are at the time of diagnoses largely predictable through a relatively limited number of features, dominated by age, BMI and comorbidities, effectively proxies for frailty.

As patients follow their disease trajectories, differential features supersede each other in prognostic importance, and prognostic models should thus consider the patient’s place in the disease trajectory.

The results of the external validation did, however, show an overall reduction in the model’s classification ability when the UK biobank cohort was analysed. This will impact on the generalizability of the presented models, but results should be interpreted with caution. As such, the UK cohort was assembled for the purpose of biobanking studies, and thus comprise a highly selected subset of patients, whereas the Danish cohort was population wide in the two analysed geographical regions. Demographic data also highlights differences in the two populations, including an age difference between groups. Actually, when predicting death for ICU patients, where demographics are similar, we do not observe a reduction in model performance.

The differences in results can be explained by the change of the underlying data distribution. The results demonstrate that caution should be exercised when evaluating whether ML models are useful for local health care practitioners if developed on other cohorts, especially when developed on early phase COVID-19 data. As such, significant variations in national factors such as isolation policies and triage for ICU and mechanical ventilation, population demographics etc. may impact on results. This notion is supported by the finding that our model retained reasonable classification ability for mortality in UK patients, but failed to predict ICU admission risk.

These results could thus indicate that potential users of ML models for COVID-19 patients should carefully examine the generalizability of the training cohort and healthcare infrastructure where patients originated from and compare these with local features prior to model usage.

Our study has several limitations. The number of patients available for this analysis was limited, and additional patient data could change the results. Secondly, we have extracted a subset of clinical variables from the EHR system. Analysing other features could affect the model. Furthermore, the changing criteria for SARS-CoV-2 testing associated with the course of the pandemic, likely also affects the results.

Even with these limitations, we may conclude that ML may be leveraged to perform outcome prediction in COVID-19 patients, as well as serve as a potential tool for identifying drivers and prognostic markers.

## Data Availability

The presented data have not been made publicly available, due to patient data safety concerns.

## Acknowledgments

The study was funded by grants from the Novo Nordisk foundation to MS (#NNF20SA0062879 and #NNF19OC0055183). Imaging for the online calculator was created by Fusion Medical Imaging.

## SUPPLEMENTARY TABLES

**Supplementary Table S1:** International Classification of Diseases (ICD) 10 and Anatomical Therapeutic Classification (ATC) codes used of the definition of chosen comorbidities. ICD- 10 codes list the presence of a given comorbidity in the Electronic Health Record, whereas the ATC code indicates that the patient is on a drug targeting the specific comorbidity. A comorbidity was classified if either the ICD-10 or ATC code was present for the given patient. COPD: Chronic Obstructive Lung Disease Neurological manifestations: Pre-existing neurodegenerative diseases such as Parkinson’s disease.

| Comorbidity | Included ICD-10 codes | Excluded ICD-10 codes | Included ATC codes | Excluded ATC codes |
| --- | --- | --- | --- | --- |
| Diabetes | E10,E11,E12,E13,E14,H28.0<br>H36.0 |  | A10 |  |
| Ischemic Heart Disease | I20, I212, I23, I24, I25 |  | CO1D |  |
| Arrhythmias | I47, I48, I49 |  | CO1AA05 |  |
| Heart Failure | I50, I42.0, I42.2, I42.6,<br>I42.7, I42.8, I42.9, I11.0,<br>I13.0, I13.2, |  | CO9DX04 |  |
| Stroke | G45, G46, I60, I62, I63, I64,<br>I65, I66, I67, I68, I69 |  | BO1AC30,<br>BO1AC07 |  |
| COPD or Asthma | J40, J41, J42, J43, J44, J45,<br>J46, J47 |  |  |  |
| Sleep apnea | G47.3 |  |  |  |
| Arthritis | MO5, MO.6 |  |  |  |
| Osteoporosis | M80, M81, M82 |  | MO5B,<br>GO3XC01,<br>HO5AA02,<br>HO5AA03 |  |
| Dementia | F00, G30, F01, F02.0, F03.9,<br>G31.8B, G31.8E, G31.9,<br>G31.0B |  | NO6D |  |
| Mental disorder | F20, F21, F22, F25, F28,<br>F29, F31 |  |  |  |
| Immunodeficiency | D80, D81, D82, D83, D84 |  |  |  |
| Neurological<br>Manifestations | G | G45, G46 |  |  |
| Cancer |  |  |  | LO1AA01,<br>LO1BA01,<br>LO1X02 |
| Chronic Kidney<br>Failure | N18 | N181 |  |  |

**Supplementary Table S2:** List of extracted laboratory tests, where available.

| **Group** | **Test** | **Abbreviation** | **Analysed from** |
| --- | --- | --- | --- |
| Inflammation/Infection | C-reactive protein | CRP | Plasma |
|  | Lymphocyte Count | - | Blood |
|  | Leucocyte Count | - | Blood |
|  | Neutrophil Count | - | Blood |
| Liver | Lactate Dehydrogenase | LDH | Plasma |
|  | Alanine<br>Aminotransferase | ALAT | Plasma |
| Coagulation and blood | D-Dimer | - | Plasma |
|  | Hemoglobin | Hgb | Blood |
|  | Ferritine | - | Plasma |
| Renal | Blood Urea Nitrogen | BUN | Plasma |
|  | Creatinine | - | Plasma |
| Arterial Blood gas | Base Excess | BE | Plasma |
|  | Bicarbonate | - | Plasma |
|  | Lactate | - | Plasma |
|  | pO<sub>2</sub> | - | Plasma |
|  | pCO<sub>2</sub> | - | Plasma |
|  | pH | - | Plasma |

**Supplementary Table S3:** List of extracted vital signs

|  |
| --- |
| Pulse |
| Temperature |
| Diastolic Blood Pressure |
| Systolic blood pressure |
| Early Warning Score |
| Oxygen supplementation (liters per minute) |
| Oxygen Saturation |

**Supplementary Table S4:** Data availability in percentages

|  | All patients | Non-hospitalized | Hospitalized patients | Hospitalized patients without ICU admission | Hospitalized patients with ICU admission | Survivors | Non-survivors |
| --- | --- | --- | --- | --- | --- | --- | --- |
| BMI | 67.9 | 63.2 | 76.9 | 74.4 | 92.8 | 66.8 | 80.9 |
| CRP (mean) | 31.1 | 1.8 | 86.9 | 86.7 | 88.4 | 26.4 | 84.6 |
| CRP (slope) | 8.9 | 0.2 | 25.4 | 19.0 | 66.9 | 7.2 | 28.1 |
| CRP (most recent) | 31.1 | 1.8 | 86.9 | 86.7 | 88.4 | 26.4 | 84.6 |
| Lymphocyte Count (mean) | 28.9 | 1.7 | 80.7 | 80.5 | 82.3 | 24.5 | 77.8 |
| Lymphocyte Count (slope) | 7.2 | 0.0 | 20.7 | 14.7 | 59.7 | 5.8 | 22.8 |
| Lymphocyte Count (most recent) | 28.9 | 1.7 | 80.6 | 80.4 | 82.3 | 24.5 | 77.8 |
| Lactate dehydrogenase (mean) | 25.7 | 1.2 | 72.3 | 71.2 | 79.0 | 21.9 | 68.8 |
| Lactate dehydrogenase (slope) | 4.4 | 0.0 | 12.6 | 7.4 | 46.4 | 3.6 | 12.7 |
| Lactate dehydrogenase (most recent) | 25.7 | 1.2 | 72.3 | 71.2 | 79.0 | 21.9 | 68.8 |
| Alanine aminotransferase (mean) | 28.8 | 1.5 | 80.8 | 80.1 | 85.1 | 24.4 | 78.1 |
| Alanine aminotransferase (slope) | 5.5 | 0.0 | 15.8 | 10.2 | 52.5 | 4.4 | 17.6 |
| Alanine aminotransferase (most recent) | 28.8 | 1.5 | 80.8 | 80.1 | 85.1 | 24.4 | 78.1 |
| Hemoglobin (mean) | 31.5 | 2.2 | 87.3 | 87.0 | 89.0 | 26.8 | 84.6 |
| Hemoglobin (slope) | 20.0 | 0.7 | 56.8 | 53.1 | 80.7 | 15.9 | 66.4 |
| Hemoglobin (most recent) | 31.5 | 2.2 | 87.3 | 87.0 | 89.0 | 26.8 | 84.6 |
| White blood cells (mean) | 31.0 | 1.9 | 86.4 | 86.1 | 88.4 | 26.2 | 84.0 |
| White blood cells (slope) | 8.8 | 0.0 | 25.4 | 18.8 | 68.0 | 7.0 | 29.9 |
| White blood cells (most recent) | 30.7 | 1.9 | 85.4 | 85.4 | 85.6 | 26.0 | 83.0 |
| Neutrophil (mean) | 29.0 | 1.7 | 80.9 | 80.6 | 82.3 | 24.6 | 78.1 |
| Neutrophil (slope) | 7.6 | 0.0 | 21.7 | 15.2 | 64.1 | 6.0 | 24.7 |
| Neutrophil (most recent) | 29.0 | 1.7 | 80.9 | 80.6 | 82.3 | 24.6 | 78.1 |
| D dimer (mean) | 7.9 | 0.2 | 22.5 | 19.7 | 40.9 | 6.6 | 21.9 |
| D dimer (slope) | 1.1 | 0.0 | 3.0 | 0.7 | 18.2 | 0.8 | 4.0 |
| D dimer (most recent) | 7.9 | 0.2 | 22.5 | 19.7 | 40.9 | 6.6 | 21.9 |
| Blood urea nitrogen (mean) | 27.5 | 1.2 | 77.6 | 76.9 | 82.3 | 23.2 | 75.6 |
| Blood urea nitrogen (slope) | 5.1 | 0.0 | 14.6 | 9.0 | 51.4 | 4.1 | 16.4 |
| Blood urea nitrogen (most recent) | 27.5 | 1.2 | 77.6 | 76.9 | 82.3 | 23.2 | 75.6 |
| Creatinine (mean) | 31.0 | 1.7 | 86.8 | 86.5 | 88.4 | 26.2 | 84.9 |
| Creatinine (slope) | 10.4 | 0.0 | 29.9 | 23.9 | 69.1 | 7.9 | 38.0 |
| Creatinine (most recent) | 31.0 | 1.7 | 86.8 | 86.5 | 88.4 | 26.2 | 84.9 |
| Ferritin (mean) | 9.3 | 0.3 | 26.4 | 23.9 | 42.5 | 7.9 | 25.6 |
| Ferritin (slope) | 1.6 | 0.0 | 4.5 | 1.8 | 22.1 | 1.2 | 5.2 |
| Ferritin (most recent) | 9.3 | 0.3 | 26.4 | 23.9 | 42.5 | 7.9 | 25.6 |
| Base excess (mean) | 19.1 | 0.9 | 53.6 | 49.5 | 80.7 | 15.2 | 62.0 |
| Base excess (slope) | 7.8 | 0.3 | 22.1 | 15.0 | 68.0 | 5.6 | 31.8 |
| Base excess (most recent) | 19.0 | 0.9 | 53.4 | 49.5 | 79.0 | 15.1 | 62.0 |
| HCO<sub>3</sub> (mean) | 19.6 | 1.0 | 55.1 | 51.0 | 81.8 | 15.6 | 64.2 |
| HCO<sub>3</sub> (slope) | 7.7 | 0.2 | 21.9 | 14.9 | 68.0 | 5.6 | 31.5 |
| HCO<sub>3</sub> (most recent) | 19.6 | 1.0 | 55.1 | 51.0 | 81.8 | 15.6 | 64.2 |
| Lactate (mean) | 14.7 | 0.9 | 41.0 | 37.6 | 63.0 | 11.8 | 47.2 |
| Lactate (slope) | 6.1 | 0.3 | 17.2 | 11.7 | 53.0 | 4.4 | 24.7 |
| Lactate (most recent) | 14.7 | 0.9 | 41.0 | 37.6 | 63.0 | 11.8 | 47.2 |
| O<sub>2</sub> (mean) | 18.2 | 0.9 | 51.2 | 46.8 | 80.1 | 14.4 | 61.1 |
| O<sub>2</sub> (slope) | 7.2 | 0.2 | 20.4 | 13.5 | 65.2 | 5.2 | 28.4 |
| O<sub>2</sub> (most recent) | 18.2 | 0.9 | 51.2 | 46.8 | 80.1 | 14.4 | 61.1 |
| pCO<sub>2</sub> (mean) | 18.2 | 0.9 | 51.3 | 46.9 | 80.1 | 14.4 | 61.1 |
| pCO<sub>2</sub> (slope) | 7.3 | 0.2 | 20.6 | 13.6 | 66.3 | 5.3 | 29.3 |
| pCO<sub>2</sub> (most recent) | 18.2 | 0.9 | 51.3 | 46.9 | 80.1 | 14.4 | 61.1 |
| ph (mean) | 18.2 | 0.9 | 51.3 | 46.9 | 80.1 | 14.4 | 61.1 |
| ph (slope) | 6.9 | 0.2 | 19.8 | 13.1 | 63.5 | 5.1 | 28.1 |
| ph (most recent) | 18.2 | 0.9 | 51.3 | 46.9 | 80.1 | 14.4 | 61.1 |
| pO<sub>2</sub> (mean) | 18.2 | 0.9 | 51.2 | 46.8 | 80.1 | 14.4 | 61.1 |
| pO<sub>2</sub> (slope) | 7.6 | 0.2 | 21.5 | 14.4 | 67.4 | 5.5 | 30.6 |
| pO<sub>2</sub> (most recent) | 18.2 | 0.9 | 51.2 | 46.8 | 80.1 | 14.4 | 61.1 |
| Pulse (mean) | 33.5 | 4.8 | 88.0 | 87.9 | 89.0 | 28.8 | 85.2 |
| Pulse (slope) | 27.8 | 0.7 | 79.4 | 78.4 | 86.2 | 23.2 | 79.6 |
| Pulse (most recent) | 33.5 | 4.8 | 88.0 | 87.9 | 89.0 | 28.8 | 85.2 |
| Temperature (mean) | 33.3 | 4.5 | 88.2 | 88.0 | 89.0 | 28.7 | 84.9 |
| Temperature (slope) | 27.2 | 0.5 | 78.0 | 77.0 | 84.5 | 22.8 | 75.9 |
| Temperature (most recent) | 33.3 | 4.5 | 88.2 | 88.0 | 89.0 | 28.7 | 84.9 |
| Early Warning Score (mean) | 29.1 | 2.1 | 80.4 | 79.4 | 86.7 | 24.3 | 82.1 |
| Early Warning Score (slope) | 23.9 | 0.3 | 68.7 | 66.5 | 82.9 | 19.5 | 72.5 |
| Early Warning Score (most recent) | 26.0 | 2.0 | 71.7 | 69.6 | 85.1 | 21.4 | 77.2 |
| Respiratory rate (mean) | 33.1 | 4.4 | 87.5 | 87.4 | 88.4 | 28.5 | 84.6 |
| Respiratory rate (slope) | 25.0 | 0.4 | 71.7 | 69.8 | 84.0 | 20.6 | 73.8 |
| Respiratory rate (most recent) | 33.1 | 4.4 | 87.5 | 87.4 | 88.4 | 28.5 | 84.6 |
| Saturation (mean) | 33.5 | 4.8 | 88.2 | 88.0 | 89.0 | 28.9 | 85.2 |
| Saturation (slope) | 25.6 | 0.7 | 72.9 | 71.1 | 84.5 | 21.2 | 74.4 |
| Saturation (most recent) | 33.5 | 4.8 | 88.2 | 88.0 | 89.0 | 28.9 | 85.2 |

**Supplementary Table S5:** Demographic information on the group SARS-CoV-2 positive patients from the United Kingdom biobank cohort used for external validation. The table presents information on the full cohort (admitted and non-admitted SARS-CoV-2 positive patients) as well as subgroups admitted to a hospital and Intensive Care Unit (ICU) respectively. Furthermore, differential demographics between survivors and non-survivors (in-hospital mortality) is presented. Continuous variables are presented as medians with (interquartile range) **p<0.001 when subgroups are compared (e.g. hospitalized vs. non-hospitalized, ICU vs. non-ICU, survivors vs. non- survivors). COPD: Chronic Obstructive Pulmonary Disease.

|  | All SARS-CoV-2 patients (n=1650) | Non-hospitalized (n=897) | Hospitalized patients (n=753) | Hospitalized patients without ICU admission (n=1519) | Hospitalized patients with ICU admission (n=131) | Survivors (n=1345) | Non-survivors (n=305) |
| --- | --- | --- | --- | --- | --- | --- | --- |
| Body Mass Index | 28.0 (25.1-31.5) | 27.3 (24.6-30.5) | 28.8 (25.9-32.6)** | 27.8 (25.0-31.2) | 29.9 (27.1-33.8)** | 27.7 (25.0-31.2) | 28.8 (25.9-32.6) |
| Age | 69.0 (59.0-76.0) | 64.0 (57.0-75.0) | 73.0 (67.0-78.0)** | 69.0 (59.0-77.0) | 69.0 (61.5-75.0) | 66.0 (58.0-75.0) | 76.0 (71.0-79.0)** |
| Male Sex | 47.8 | 56.6 | 37.2** | 49.0 | 32.8** | 50.7 | 34.8** |
| Diabetes | 14.1 | 10.3 | 18.7** | 14.3 | 12.2 | 12.1 | 23.0** |
| Ischemic heart disease | 14.7 | 10.6 | 19.7** | 14.8 | 13.7 | 12.6 | 24.3** |
| Heart failure | 4.2 | 2.7 | 6.0** | 4.2 | 3.8 | 3.4 | 7.5** |
| Arrhythmia | 9.9 | 6.8 | 13.5** | 10.0 | 8.4 | 8.4 | 16.4** |
| Stroke | 5.9 | 5.8 | 6.1 | 6.3 | 1.5 | 5.0 | 10.2** |
| COPD or Asthma | 15.2 | 12.2 | 18.7** | 15.1 | 15.3 | 13.8 | 21.0** |
| Sleep apnoea | 0.0 | 0.0 | 0.0 | 0.0 | 0.0 | 0.0 | 0.0 |
| Arthritis | 2.4 | 1.6 | 3.5 | 2.4 | 3.1 | 1.8 | 5.2** |
| Osteoporosis | 3.0 | 2.5 | 3.7 | 3.1 | 2.3 | 2.8 | 3.9 |
| Dementia | 2.1 | 2.5 | 1.7 | 2.3 | 0.0 | 1.6 | 4.3 |
| Severe mental disorder | 0.8 | 0.7 | 1.1 | 0.8 | 1.5 | 0.6 | 2.0 |
| Immunodeficiencies | 0.3 | 0.3 | 0.3 | 0.3 | 0.0 | 0.2 | 0.7 |
| Neurological manifestations | 19.2 | 16.6 | 22.3 | 19.6 | 14.5 | 17.2 | 27.9** |
| Cancer | 12.8 | 10.3 | 15.9** | 12.9 | 12.2 | 11.4 | 19.3** |
| Chronic | 5.2 | 3.7 | 7.0 | 5.1 | 6.1 | 4.6 | 7.9 |
| kidney failure |  |  |  |  |  |  |  |
| Dialysis | 0.9 | 1.1 | 0.7 | 0.9 | 0.8 | 1.0 | 0.7 |
| Hypertension | 35.5 | 28.2 | 44.1** | 35.3 | 37.4 | 31.1 | 54.8** |

**Supplementary Table S6:** Main results from the external validation of the prediction models. Performance metrics are presented as the Receiver Operating Characteristics Area Under the Curve (ROC-AUC) for True/False positive rates (TPR/FPR) and Precision/Recall (Pre/Rec). *Model is significantly (p<0.01) better than the base prediction model (Age+gender+Body Mass Index, BMI) --: Insufficient data available at the time point, or prediction irrelevant (e.g. predicting hospital admission for patients already in the ICU).

|  | Hospital admission |  | ICU admission |  | Death |  |
| --- | --- | --- | --- | --- | --- | --- |
|  | TPR/FPR | Pre/Rec | TPR/FPR | Pre/Rec | TPR/FPR | Pre/Rec |
| Diagnoses |  |  |  |  |  |  |
| Age+Gender+BMI | 0.648 | 0.561 | 0.509 | 0.083 | 0.716 | 0.294 |
| +Comorbidities | 0.664* | 0.58 | 0.548 | 0.094 | 0.724 | 0.302 |
| Admission |  |  |  |  |  |  |
| Age+Gender+BMI | -- | -- | 0.557 | 0.189 | 0.607 | 0.394 |
| +Comorbidities | -- | -- | 0.518 | 0.182 | 0.625 | 0.413 |
| Pre-ICU |  |  |  |  |  |  |
| Age+Gender+BMI | -- | -- | -- | -- | 0.623 | 0.524 |
| +Comorbidities | -- | -- | -- | -- | 0.679 | 0.606 |
| Post-ICU |  |  |  |  |  |  |
| Age+Gender+BMI | -- | -- | -- | -- | 0.623 | 0.524 |
| +Comorbidities | -- | -- | -- | -- | 0.674 | 0.58 |

## SUPPLEMENTARY FIGURES

**Supplementary Figure S1:**
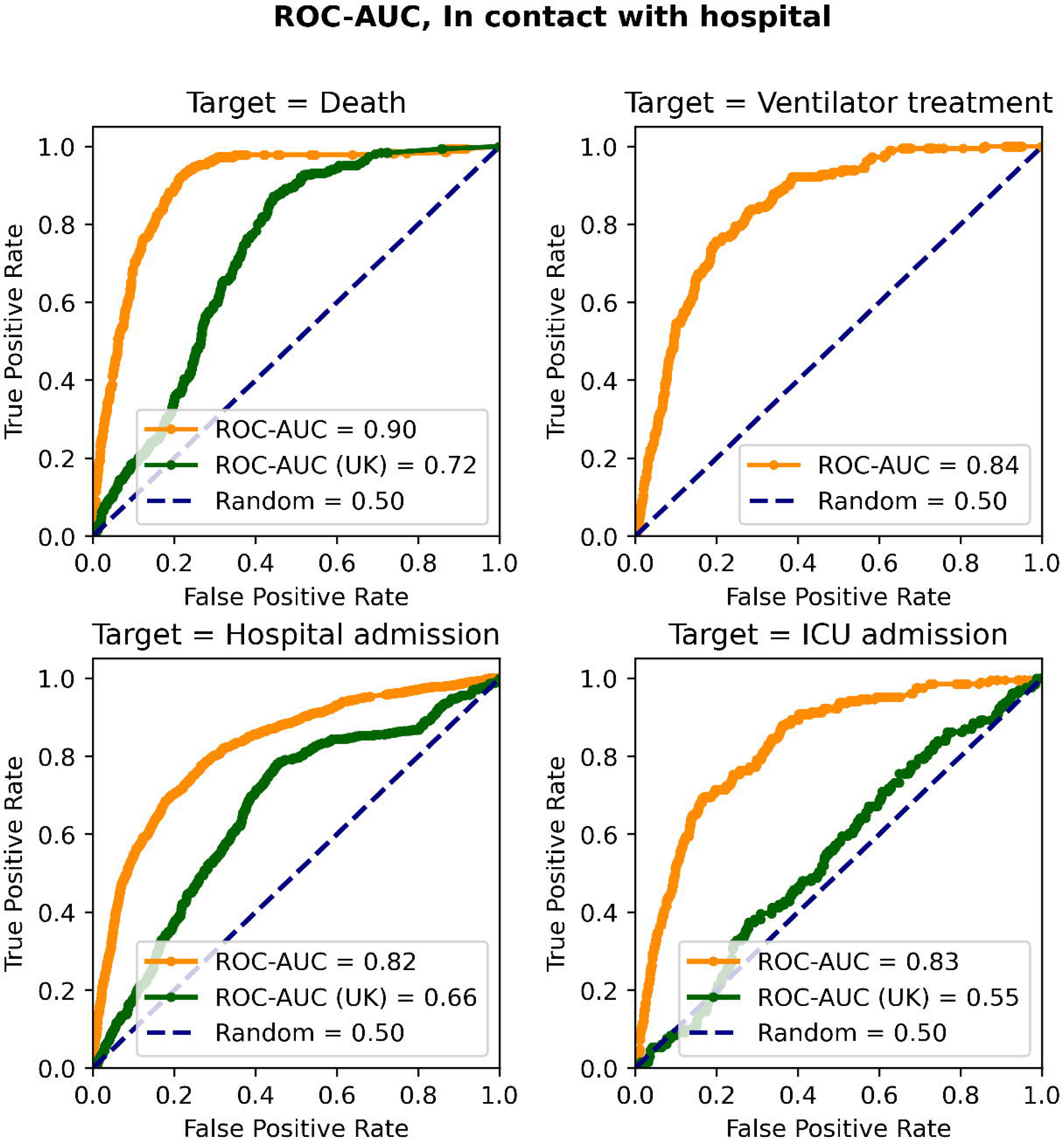
Receiver operator characteristics – area under the curve (ROC-AUC) of the patients tested positive for SARS-CoV-2 (Diagnosis model). The model predicted risks of hospital admission (bottom left), intensive care admission (bottom right), ventilator treatment (top right) and death (top left). Orange line indicates results obtained on Danish data; green line indicates results obtained in external validation data from the UK biobank.

**Supplementary Figure S2:**
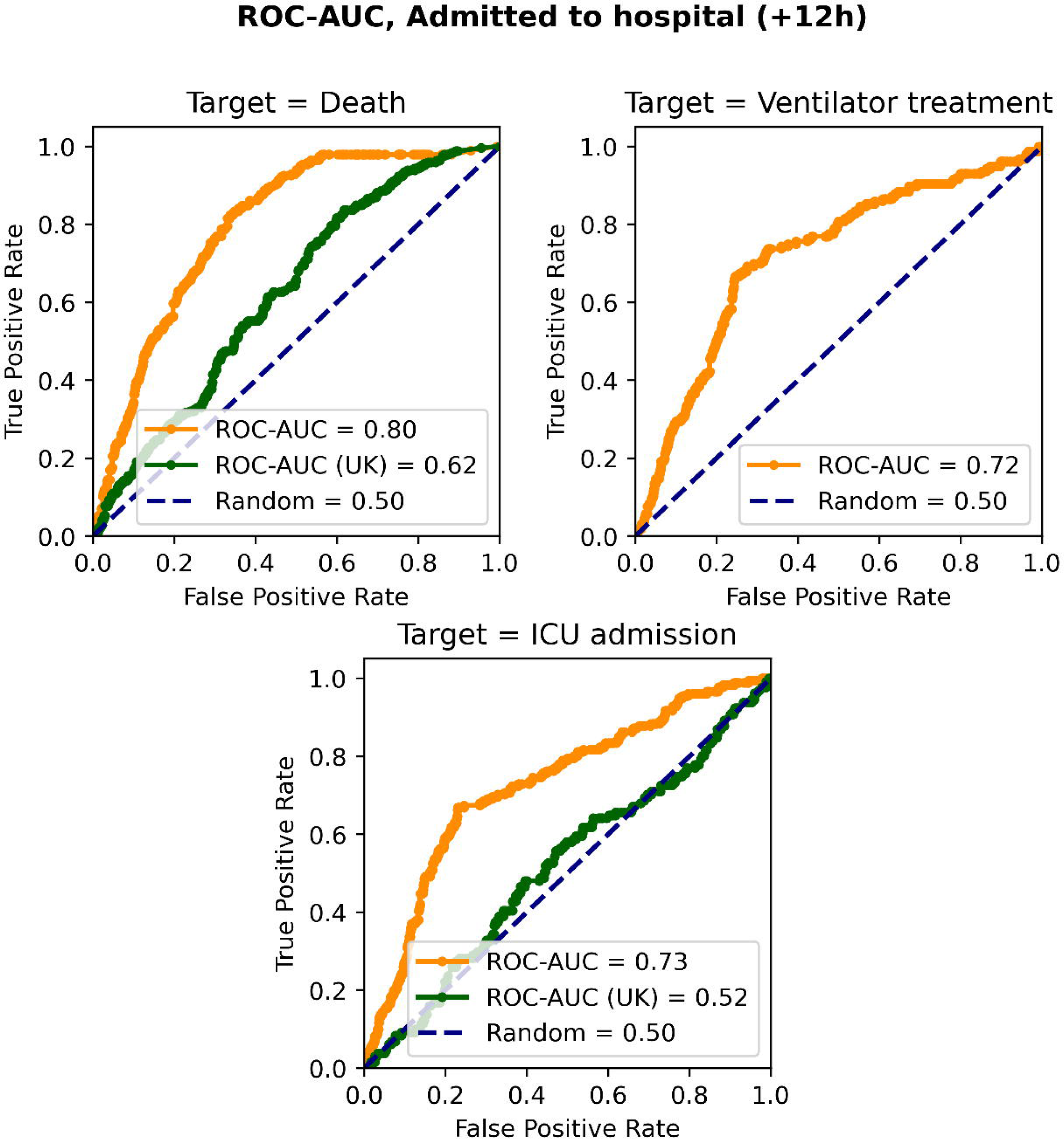
Receiver operator characteristics – area under the curve (ROC-AUC) of the patients admitted to the hospital. Model inputs included data available up to 12 hours after the admission timepoint (admission model). The model predicted risks of intensive care admission (bottom), ventilator treatment (top right) and death (top left). Orange line indicates results obtained on Danish data; green line indicates results obtained in external validation data from the UK biobank.

**Supplementary Figure S3:**
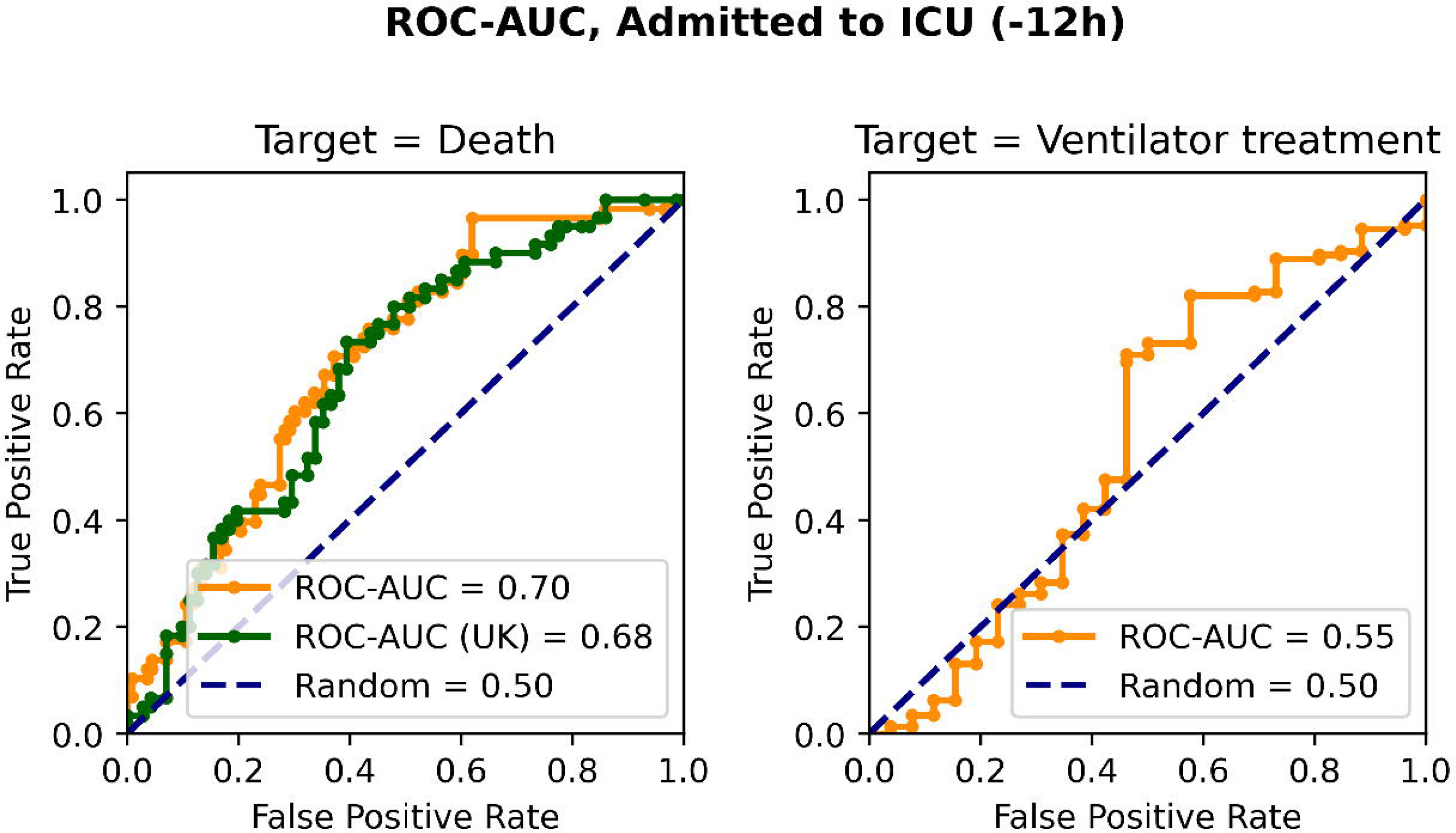
Receiver operator characteristics – area under the curve (ROC-AUC) of the patients admitted to the Intensive Care Unit (ICU). Model inputs included data available from the admission model, as well as temporal features available during the 12 hours leading up to ICU admission. The model predicted risks of ventilator treatment (right) and death (left). Orange line indicates results obtained on Danish data; green line indicates results obtained in external validation data from the UK biobank.

**Supplementary Figure S4:**
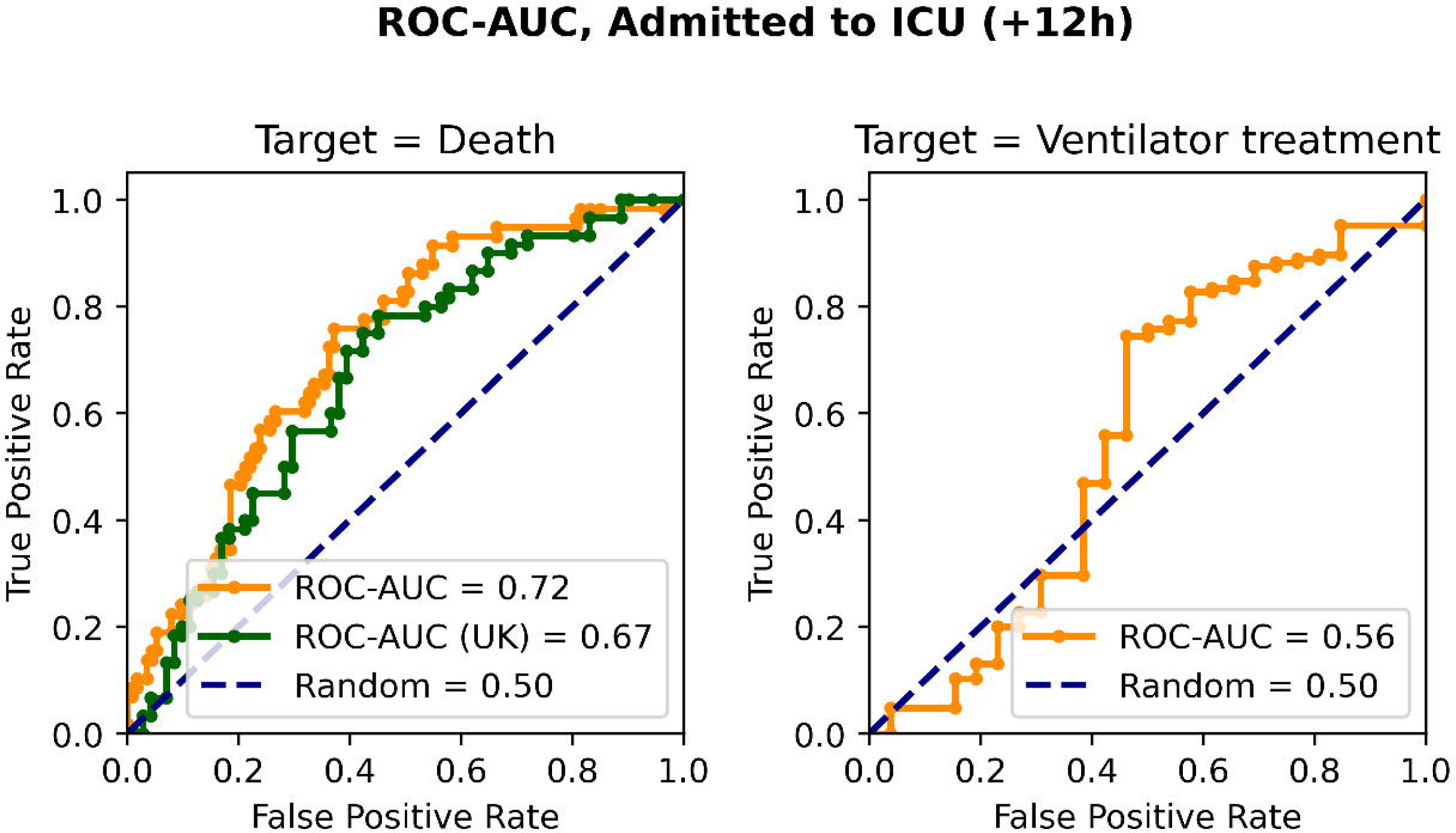
Receiver operator characteristics – area under the curve (ROC-AUC) of the patients admitted to the Intensive Care Unit (ICU). Model inputs included data available from the admission model, as well as temporal features available during the 12 hours after ICU admission. The model predicted risks of ventilator treatment (right) and death (left). Orange line indicates results obtained on Danish data; green line indicates results obtained in external validation data from the UK biobank.

**Supplementary Figure S5:**
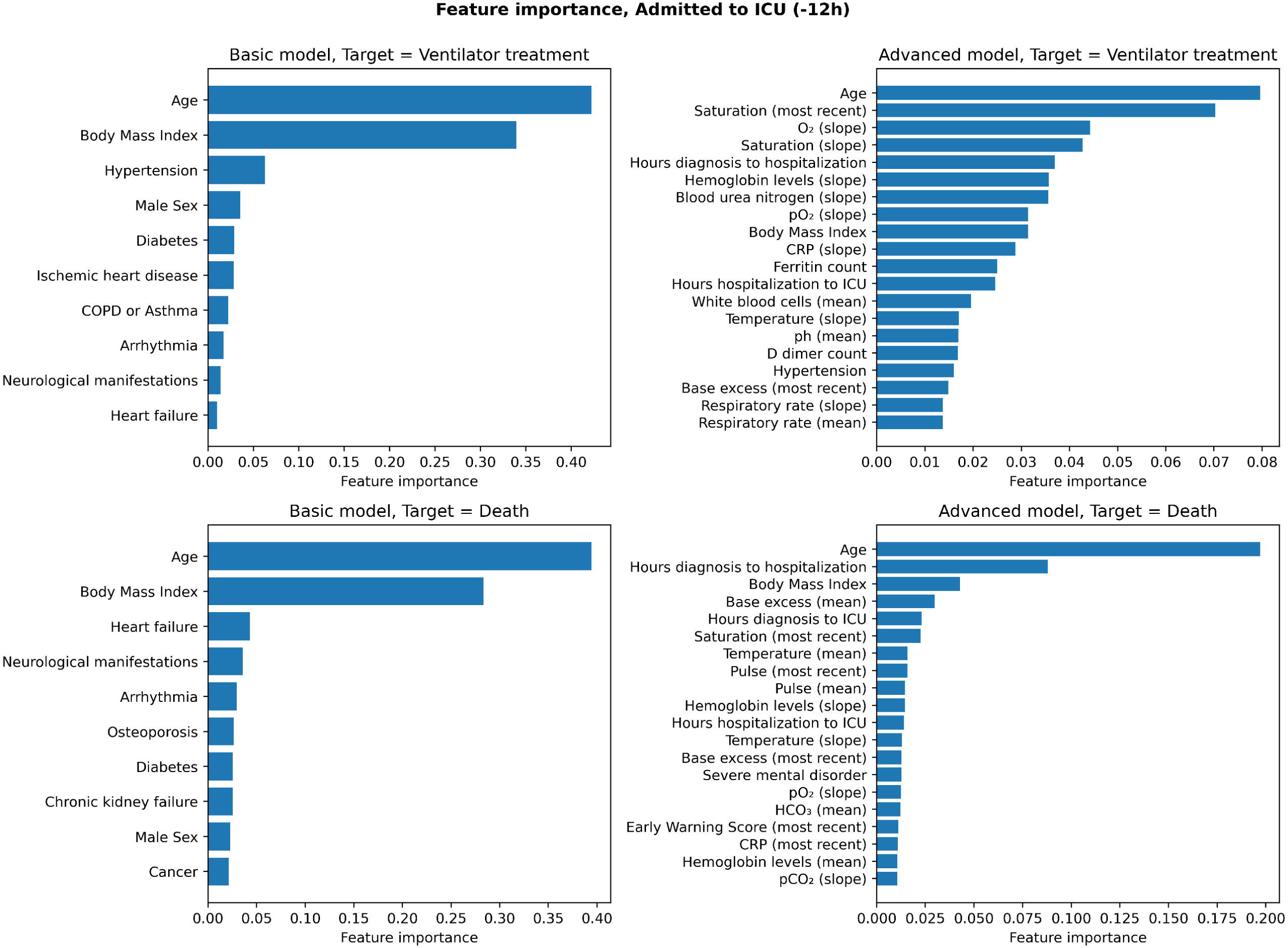
Feature importance for the basic (including age, sex and body mass index) and advanced (all data) pre-ICU model, predicting risk ventilator treatment (first row) and death (second row).

**Supplementary Figure S6:**
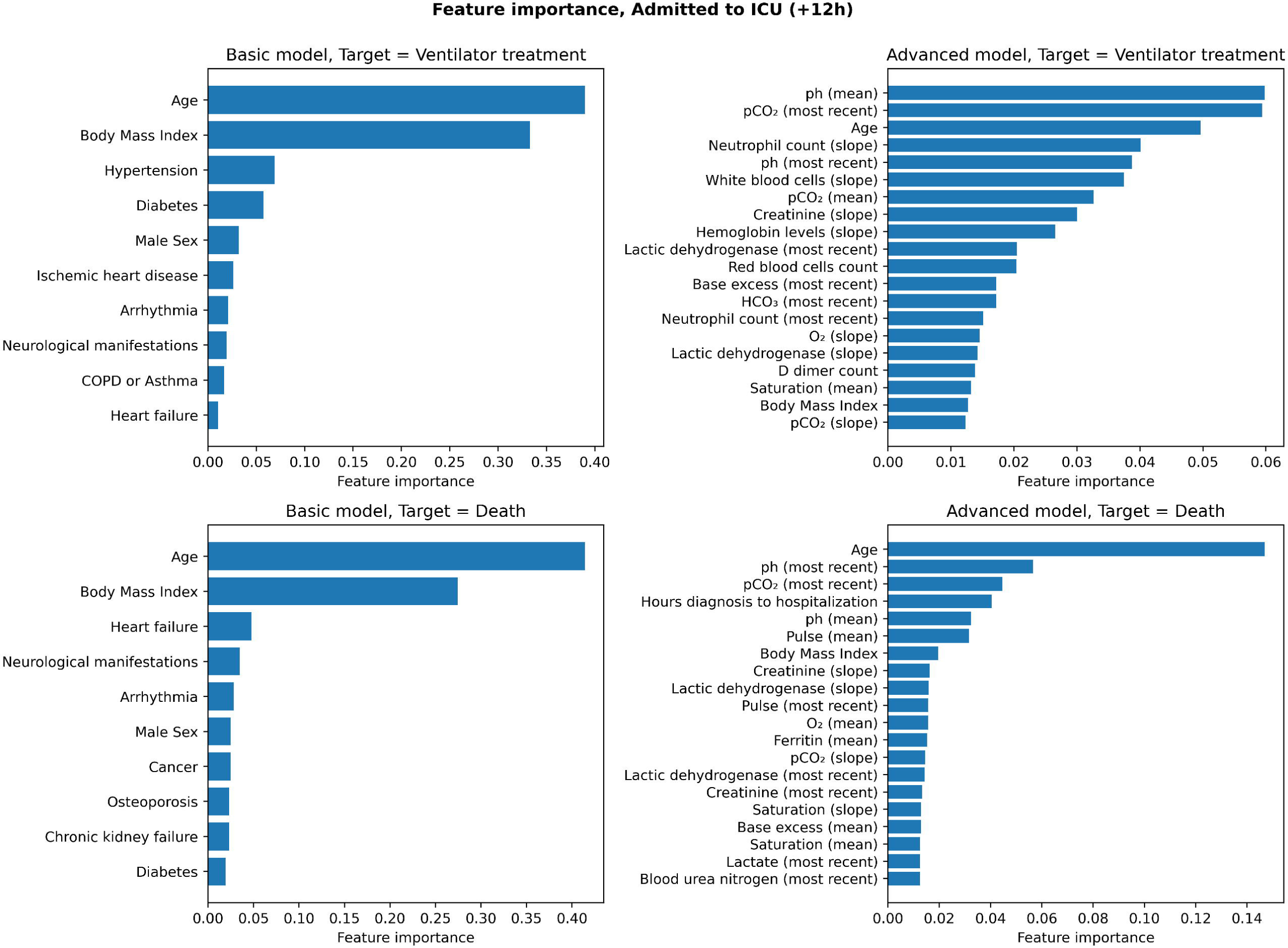
Feature importance for the basic (including age, sex and body mass index) and advanced (all data) post-ICU model, predicting risk ventilator treatment (first row) and death (second row).

**Supplementary Figure S7:**
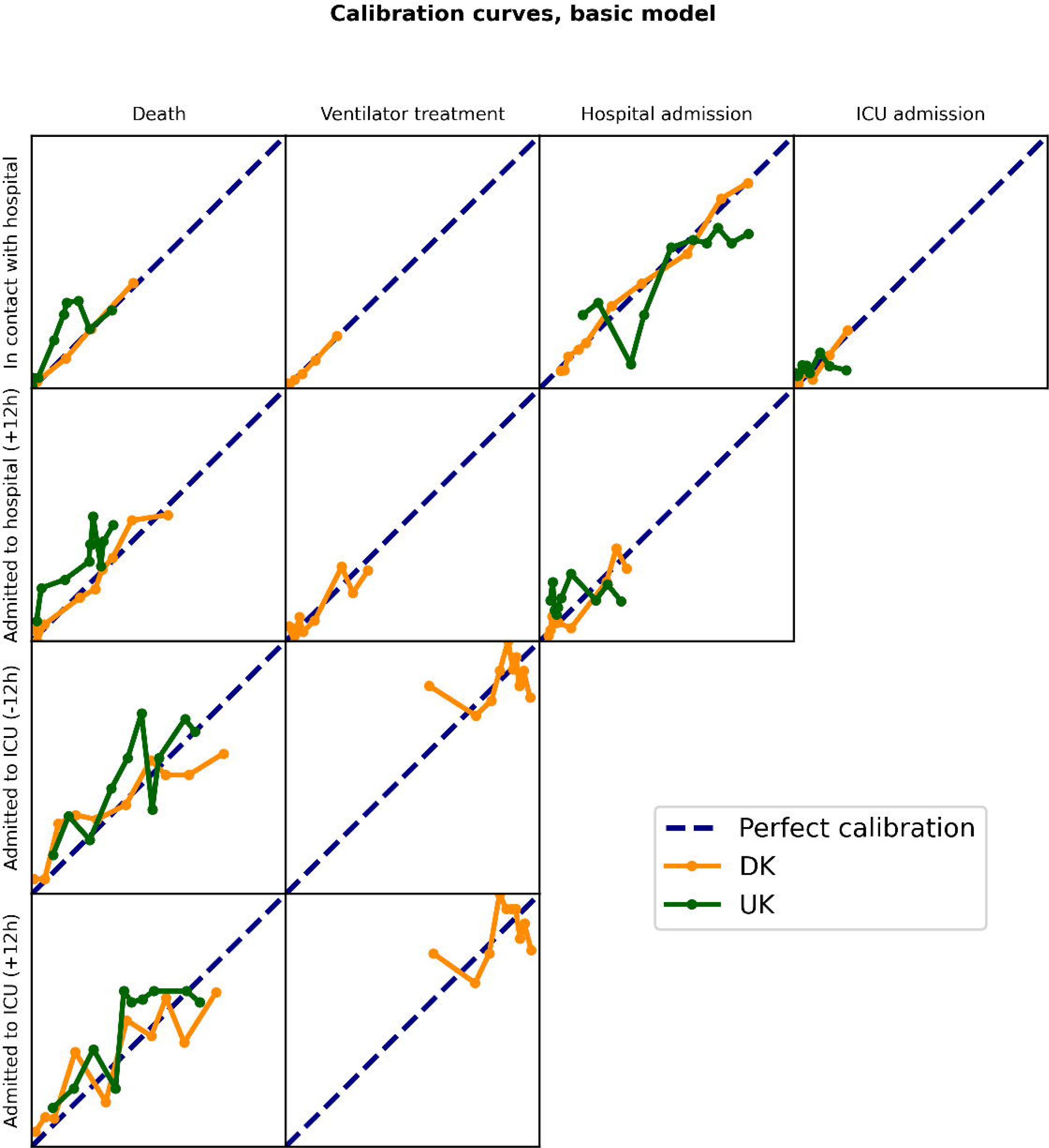
Calibration curves for basic models. Orange line indicates Danish data, green line indicates data from the United Kingdom biobank. For each calibration plot, the abscissa shows the mean of the predictions within a bin and the ordinate shows the fraction of positive labels for the corresponding samples.

**Supplementary Figure S8:**
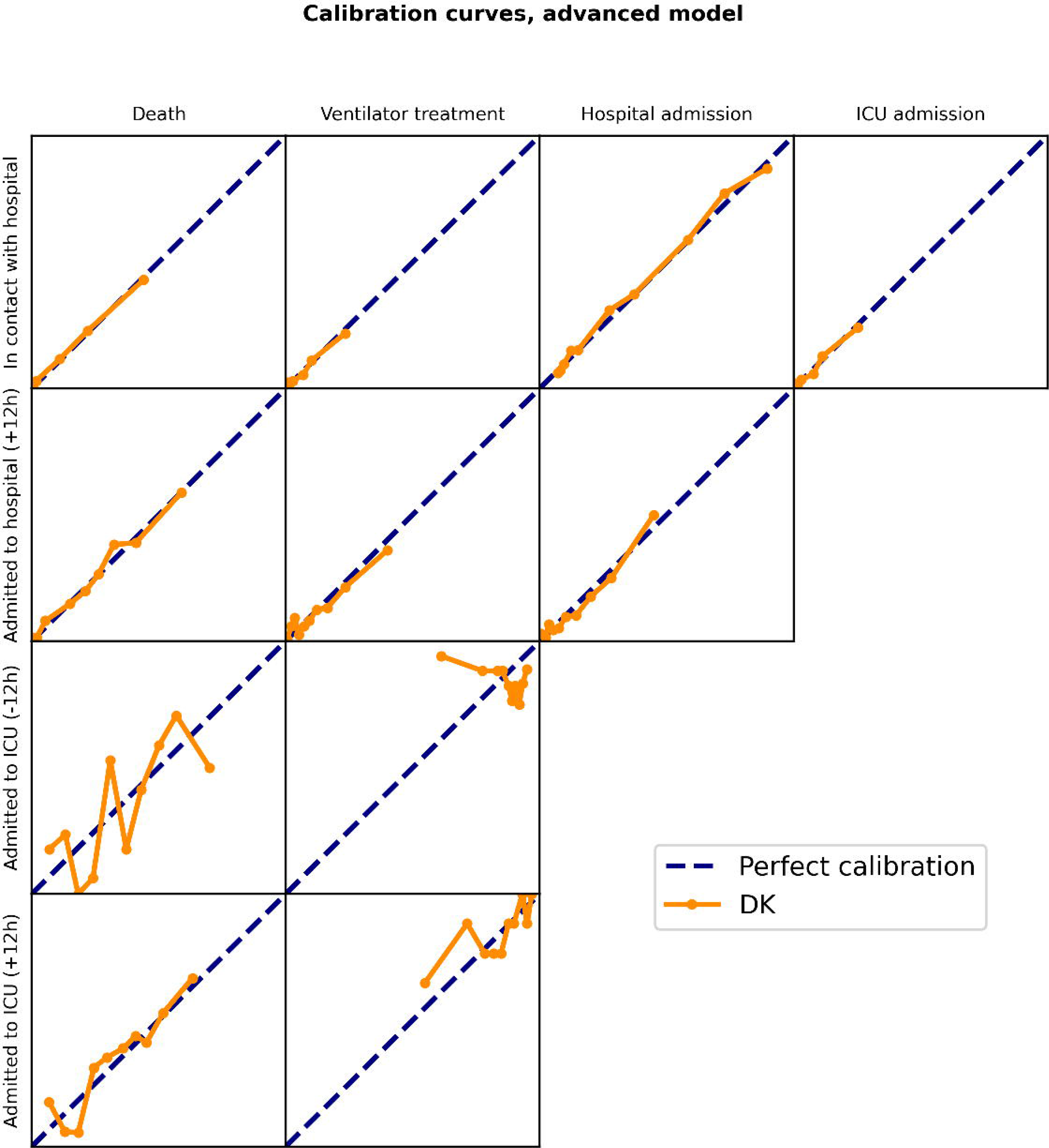
Calibration curves for advanced models. Orange line indicates Danish data. For each calibration plot, the abscissa shows the mean of the predictions within a bin and the ordinate shows the fraction of positive labels for the corresponding samples.

## Notes

### Competing Interest Statement

The authors have declared no competing interest.

### Author Declarations

As per your request for resubmission, we have expanded the information in the manuscript as detailed below. Under Danish Law, ethical and legal approval for access to patient charts for research purposes is governed by the Danish Patients Safety Authority (Styrelsen for Patientsikkerhed, en.stps.dk). Storage and handling of the data is approved by the Danish Data Protection Agency (Datatilsynet, www.datatilsynet.dk). Legal clearance for the study was furthermore obtained from the Danish Capital Region. Under Danish Law, Ethics committee approval (in Denmark, Videnskabsetisk komite) is required for studies requiring interaction with patients, but not for studies solely needing patient chart review and data extraction (without patient contact). For these studies, the Danish Patients Safety Authority is the ethics and legal governing body as stated. As such, the study obtained all necessary legal and ethics approval prior to commencement. New manuscript text: The study was approved by the relevant legal and ethics boards. These included the Danish Patient Safety Authority (Styrelsen for Patientsikkerhed, approval #31-1521-257) and the Danish Data Protection Agency (Datatilsynet, approval #P-2020-320) as well as the UK Biobank (Application ID #60861) COVID-19 cohort. Under Danish law, approval from these agencies are required for access to and handling of patient sensitive data, including EHR records. Legal approval for the study was furthermore obtained from the Danish Capital Region (Region Hovedstaden).

